# Health-service costs for the treatment of multidrug-resistant/rifampicin-resistant tuberculosis in children in South Africa: application of a real-world dataset

**DOI:** 10.1101/2025.06.15.25328393

**Authors:** Thomas Wilkinson, Arne von Delft, Anneke C. Hesseling, Edina Sinanovic, H. Simon Schaaf, James A. Seddon

## Abstract

**Background:** Children with multidrug-resistant (MDR) / rifampicin-resistant (RR)-tuberculosis (TB) are an important but neglected group in cost-effectiveness research. Digital health information systems enable new approaches to health-service cost analysis. The Provincial Health Data Centre (PHDC) in the Western Cape, South Africa, collates disparate health system data including hospital inpatient and outpatient data, medication, laboratory tests, and primary healthcare utilisation.

**Methods:** Cost analysis was conducted using anonymised, integrated PHDC data for children with MDR/RR-TB between 1 January 2018 and 31 December 2021. Health-service costs were assessed by patient and disease characteristics including age, sex, drug susceptibility type, site of disease, and HIV status.

**Results:** There was significant cost variation across the n=271 children in the data sample (median US$7,576 (IQR $2,725 - $22,986)). The distribution of total per patient costs fitted a gamma distribution (mean US$13,435, α = 0.93, β = 14,496). Regression analysis indicates age, disease site, living with HIV, and treatment duration had significant impact on costs, whereas the impact of resistance profile and sex was not significant.

**Conclusion:** Treatment for MDR/RR-TB in children remains costly for health systems. Utilising routinely collected, real-world data from an established health information system enables accurate and representative insights to overall costs and major cost drivers. Costs were highly skewed, with a small proportion of patients incurring very high costs. This cost analysis can assist in decision-making and programme development at local and international levels and as an input to secondary analysis.

**Key points:**

- Health-service costs of treating children who have drug resistant tuberculosis can be high, but the cost vary widely among patients
- Key factors that affect costs are age, the site of the tuberculosis disease, whether the child has HIV, and the length of time that a child receives treatment.
- Treating children in hospital is a major component of costs, indicating that programs or treatments that can reduce hospitalisation are likely to reduce costs.

## INTRODUCTION

Multidrug-resistant and rifampicin-resistant tuberculosis (MDR/RR-TB) in children is a complex and under-investigated public health issue, particularly related to costs and economic considerations for health systems. For the purposes of this analysis, MDR/RR-TB refers to disease due to *Mycobacterium tuberculosis* that is resistant to isoniazid and rifampicin (MDR-TB), or where only rifampicin resistance is established and isoniazid susceptibility is either established or unknown (RR-TB) (1). Globally, an estimated 1.3 million children developed TB disease in 2023, and more than 30,000 of these are estimated to have had MDR/RR-TB (1)(2). Health-service costs are those from the perspective of the health system or national disease programmes and exclude indirect and out-of-pocket payments by patients. Health-service costs associated with the management of any health condition is an essential input to budgeting, planning and resource allocation decisions. The lack of meaningful and localised health-service costing evidence can be a barrier to the introduction of innovations or improved approaches to delivering health care, limiting the ability of policy makers to determine how best to invest limited resources to maximise health outcomes (3). Importantly, health-service costing involves more than broad approximations of average expenditure and health-service resource utilisation. It requires a comprehensive and systematic analysis of the factors that make up major cost categories, and how these factors impact the distribution of costs among different patients (4).

Significant progress in managing childhood MDR/RR-TB, particularly a shift toward shorter, all-oral regimens, is expected to improve treatment outcomes and the overall experience of both children and their caregivers when accessing TB care. These developments will impact the costs of treatment and prevention, requiring investment in some areas while releasing health system savings in other areas. However, without precise and representative cost data, policy makers cannot make informed decisions regarding the allocation of limited National TB Programme budgets and donor funding, stalling the translation of scientific and clinical advances into meaningful outcomes for patients.

The costs to National TB Programmes associated with the management of MDR/RR-TB in children is highly uncertain and variable. This is due to differences in presentation and progression of disease, approach and response to treatment and models of care, and heterogeneity within a cohort of patients that range from newborn to 14 years of age. The recommended treatment approach will vary based on the age of the patient, medicine resistance profile, severity of disease, and site of disease (5). Treatment ranges from the delivery of standardised drug regimens in primary healthcare (PHC)/outpatient settings, to long-term individualised management in tertiary hospitals or specialised-TB hospitals (6). In addition, other localised factors including country TB programme guidance, established referral and management systems, physician judgement, and healthcare facility and healthcare worker availability will influence the disease management approach and costs of care (7). Cost analysis is also often limited by health system factors such as difficulties in collecting and coordinating available expenditure and utilisation data.

The concentration of childhood TB in low- and middle-income-country (LMIC) settings means that the ability to comprehensively identify and analyse health-service costs is often limited due to a lack of comprehensive digital health information and expenditure tracking systems. As a result, typical approaches to TB costing include patient surveys, adjunct costing within clinical trials, or manual data extraction of cost data from a limited sample of paper-based medical notes. These approaches provide valuable insights into costs of care but are limited by available resources and introduce uncertainties associated with representativeness, sampling and distribution of observed costs (8).

As children with MDR/RR-TB represent a minority of the total TB burden in local settings, there is an existing gap in the literature for costing analysis for this group. A systematic review of TB costing studies in 2020 identified 103 papers reporting costs of TB care, however, it did not identify any cost analysis of MDR/RR-TB specifically in children (9). Dodd et al. (2022) modelled the global impact and cost-effectiveness of household contact management for children at risk of MDR/RR-TB, using a normative costing approach with approximations of MDR/RR-TB costs from secondary sources (10). Some insights relating to the costs of childhood MDR/RR-TB can be inferred by analyses in adults with MDR/RR-TB or children with drug-susceptible TB (DS-TB). For example, Cox et al. (2015) assessed the cost of treating RR-TB in adults in a community-based programme in Khayelitsha, the Western Cape province of South Africa (11). Cox et al. reported a median cost per adult patient of US$5,054, with an extremely wide range between US$260 - $87,140, with higher median costs for those with outcomes recorded as treatment failure ($13,010), and lower median costs for those who died ($4,669) or who were lost to follow up (LTFU), ($2,732). Madan et al. (2020) conducted an empirical cost-effectiveness analysis of MDR-TB treatment within the STREAM trial (12). The analysis found the cost per adult patient in South Africa was $8,341 and $6,618 for long and short MDR/RR-TB treatment regimens, respectively, but was limited to the 47 adults in the trial, without differentiation by treatment outcome. Mandalakas et al. (2013) estimated the health-service costs of childhood TB in South Africa as an input to cost-effectiveness analysis (13), Applying a normative costing approach using secondary data, the analysis estimated the cost for six months of uncomplicated pulmonary DS-TB treatment as US$ 908 for children aged 0-2 years old and US$ 1,359 for children aged 3-5 years old. As a secondary analysis, the study also did not differentiate costs by treatment outcome.

The wide variation in the limited literature available demonstrates that there is currently an acute and unmet need for empirical, representative and localised health-service cost analysis in childhood MDR/RR-TB.

## METHODS

### Study design and data source

A retrospective cohort costing and regression analysis was conducted using health-service utilisation data from the Provincial Health Data Centre (PHDC) (14). The PHDC is maintained by the Department of Health and Wellness in the Western Cape province of South Africa and is a unique data repository representing 22 government health system information systems anchored to a unique patient identifier. The PHDC includes data on hospital admissions and procedures, outpatient activity, primary care activity, medicine use, diagnostics, and relevant disease registries, including South Africa’s national electronic drug-resistant TB register (EDRWeb) (15). Its primary goal is to enhance clinical care by ensuring data availability to clinicians and analysts. This is achieved by consolidating relevant data for approximately 15 million patient contacts annually at 52 hospitals and 354 PHC clinics managed by the Western Cape Provincial Department of Health and Wellness and City of Cape Town municipality (14).

For this analysis, utilisation data on all children aged from birth to 14 years old with any cross-database evidence for MDR/RR-TB between 1 January 2018 and 31 December 2021 were extracted from the PHDC. To reflect per patient costs across the entire episode of care, patients with a TB outcome of, ‘Transferred out’, or ‘Unknown/incomplete data’ were excluded from per patient costing analysis. The remaining patients had a recorded TB treatment outcome of ‘Treatment success’, ‘Unsuccessful treatment’, ‘Lost to follow up (LTFU)’, or ‘Died’ and were included in the analysis.

### Data cleaning and categorisation

Patients were stratified by reported age, sex, drug resistance profile, site of disease and HIV status. Age at TB diagnosis was used to assign the following age brackets: birth to under 2 years (i.e., 0-1 year); 2 years to under 5 years (2-4 years); and 5 years to under 15 (5-14 years) years. Consolidation of the 0-4-year age range was used for additional analysis (S1 Supplementary Appendix). These age ranges were applied based on author judgment reflecting expected parameter requirements for secondary cost effectiveness analysis of interventions for the prevention and treatment of childhood MDR/RR-TB.

To ensure that the analysis captured TB relevant events within a reasonable timeframe the analysis excluded hospital events, clinic visits, outpatient visits, procedures and laboratory tests that occurred more than 30 days before the date of the first TB-related data point or more than 30 days after the TB outcome was recorded. Events outside this period were only included in the analysis if the primary or subsidiary International Classification of Diseases 10th Revision (ICD-10) code indicated the event was TB related.

TB disease with drug resistance profiles recorded at the time of data extraction were as follows: RR-TB (rifampicin resistance confirmed without availability of drug susceptibility testing (DST) for isoniazid or other drugs, or rifampicin resistance confirmed and susceptible to INH), MDR-TB, and XDR-TB (extensively drug-resistant TB disease; i.e., MDR/RR plus fluoroquinolone and second-line injectable agent resistance).The categories of RR-TB and MDR-TB were combined within this analysis, due to a common approach to clinical management and the limited need for differentiated cost parameters in secondary analysis.

Pre-XDR-TB is TB caused by *M. tuberculosis* that fulfilled the definition of MDR/RR-TB, plus resistance to the fluoroquinolone class of drugs which are used commonly in MDR/RR-TB treatment regimens (16). In this analysis, a pre-XDR-TB category was assigned to patients initially categorised as MDR-TB within the dataset and who received no more than one prescription of a fluoroquinolone under an assumption that a fluroquinolone would have been dispensed for all MDR/RR-TB patients unless resistance was present. A resistance category “mono” was identified in the initial cohort extracted which represents patients with resistance to isoniazid and documented susceptibility to rifampicin. Patients with resistance classification of “mono” were subsequently removed from the cohort as the treatment of isoniazid mono-resistant TB is similar to drug-susceptible TB management (17).

In South Africa, children with TB disease may be initiated on treatment in an outpatient setting or in hospital (in the case of an acute admission). Patients will continue to receive their MDR/RR-TB treatment through a combination of outpatient and PHC clinic appointments (with intermittent acute hospital admissions when needed) or as an inpatient in one of the specialist TB hospitals. Patients are mainly hospitalised if long-term clinical management is required or due to social circumstances. A 2016-2017 analysis of children admitted to Brooklyn Chest Hospital (a specialist TB hospital) in the Western Cape, South Africa, found that reasons for admission were social or caregiver related (45.3%), drug-resistant TB (43.3%), TB meningitis (32.7%) and other forms of severe TB (24.0%) with 41.8% of patients with more than 1 reason for admission (18).

For this analysis, children’s encounters with the health system were categorised into four types of visits: 1) PHC clinic visits, 2) Outpatient (OP) visits, 3) Hospital Admissions to general hospitals (Hospital: General), and 4) Hospital Admissions to specialist TB hospitals (Hospital: specialist TB). The site of disease classifications (pulmonary, extrapulmonary, or unknown/other) were consolidated from the dataset (see S1 appendix for further detail). Due to the infection-risk perspective approach to recording site of disease at the point of data capture, the category of “pulmonary” may include some patients who have both pulmonary and extra-pulmonary disease, whereas the classification of “extra-pulmonary” is limited to patients who have only extra-pulmonary disease.

### Costing methodology

Utilisation data obtained from the PHDC was multiplied by local unit prices to reflect per patient health-service costs. The Uniform Patient Fee Schedule (UPFS) (2021) was used for major outpatient and hospital-related costs (e.g., day-rates, surgery, and imaging) (19). The UPFS is the charging mechanism for public sector facilities in South Africa and is developed on a cost-recovery basis. It was therefore considered to be a reasonable approximation of the cost of specific units of health care utilisation at Western Cape health facilities. The UPFS cost per inpatient stay consists of two components: a facility fee (reflecting overhead costs for the facility) and a professional fee (reflecting the costs of the healthcare professionals delivering the service such as general practitioner, specialist, nurse practitioner or allied health professional). Laboratory costs were sourced from the 2021 National Health Laboratory Service catalogue (20).

The representation of multiple databases in the PHDC enabled missing or incomplete data to be cross-checked and synthesized. Medicine usage reported in the PHDC utilises the Western Cape health facility prescribing system (JAC prescribing system). However, prescribing data in the dataset was commonly missing or incomplete for medicines dispensed from PHC clinics or ward stock. Therefore, the expected regimen based on age range and resistance profile were identified based on South African National TB programme recommendations (21) and confirmed by expert opinion as the most likely regimen for children at Western Cape health facilities during the data extraction period. Treatment duration (time from treatment initiation to recording of TB outcome) was used to calculate overall medicine costs, applying weight-adjusted cost per patient month for paediatric MDR/RR-TB drug regimens as reported by Wilkinson et al. 2024 (22) adjusted for TB-medicine pricing in 2021 using the South African National Master Health Products List (MHPL) (23). Medicine costs were also adjusted to reflect the high rate of dispensing of the oral liquid formulation of linezolid in the dataset. See S1 Supplementary appendix for further description of costing methodology, unit prices, and TB medicine cost calculations. All prices in South African Rand are converted to United Sates dollars (US$) using World Bank 2021 exchange rate.

### Analysis

Descriptive statistics were conducted for relevant study outcomes. A standard log-level regression model was specified as follows:

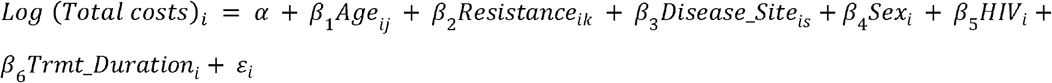

Where:

- *Total costs* is the sum of all costs incurred per patient *i*
- *Age* determines if patient *i* is in ***j*** age cohort of 0-1 years old; 2-4 years old; or 5-14 years old
- *Resistance* determines if patient *i* is in *k* drug resistance classification of either MDR/RR-TB;; Pre-XDR-TB; or XDR-TB
- *Disease_Site* determines if patient *i* is in *s* classification of either pulmonary disease, extra-pulmonary disease or “other or unknown”
- *Sex* determines if patient *i* is male (=1) or female (=0)
- *HIV* determines if patient *i* is HIV positive (=1) or HIV negative =0
- *Trmt_Duration* is a continuous variable determining number of months of treatment for patient *i*

The distribution of total costs was specified for the total cohort, and by age and resistance profile. Small patient numbers limited the reliable specification of a cost distribution for patients classified as pre-XDR-TB or XDR-TB (see S1 supplementary appendix).

The study gained ethics approval from the Stellenbosch University Health Research Ethics Committee (no. N20/09/102) and approval from the Western Cape Department of Health and Wellness (reference: WC_202109_004).

## RESULTS

The initial cohort included n=491 patients and represented more than 1.1 million data points. Excluding entries where treatment outcomes were transferred out, unknown/incomplete, or classified as isoniazid mono-resistant TB, resulted in n=271 children included in the analysis. Table 1 details key characteristics including age cohort, sex, TB drug susceptibility type, HIV status, site of TB disease, outcome, and treatment duration. The cohort was balanced between males and females and there was reasonable representation across the age cohorts: 40% of the sample was in the 5-14 years old age group, and 31% and 29% of the cohort was in the 0-1 years and 2-4 years age groups, respectively. Children living with HIV was 12% of the cohort.

**Table 1:**
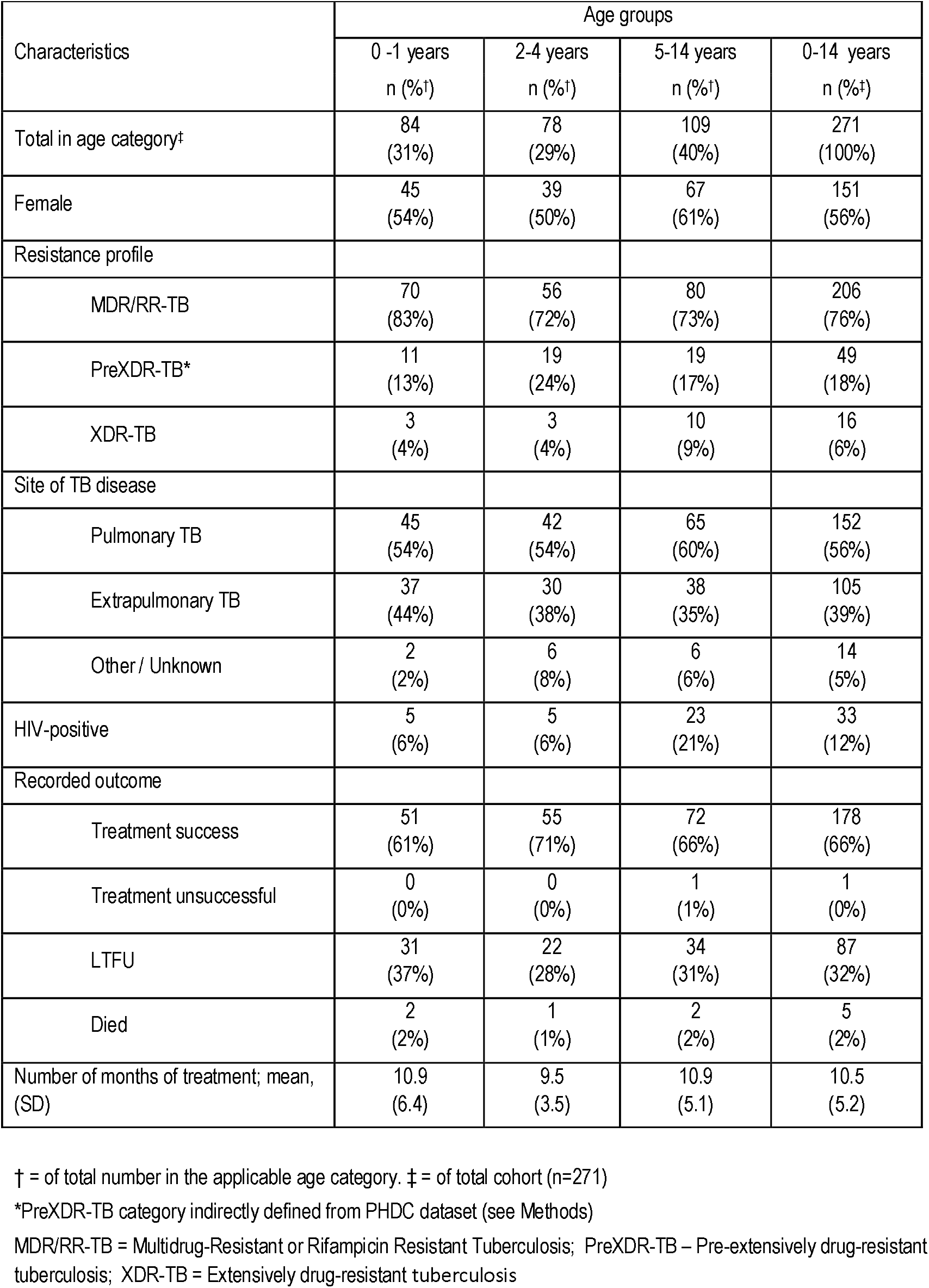
Children with multidrug-resistant/rifampicin-resistant tuberculosis demographic and clinical characteristics by age category (n=271)

The median costs of treatment by category and cost type are shown in Table 2 and Figure 1. The median total cost of treatment across the cohort was $7,576 (interquartile range (IQR) $2,725 - $22,986). Substantial differences were observed in median costs between extrapulmonary vs pulmonary disease ($14,534 vs $4,423), and younger age. Median treatment costs in the 0-1 years old age group was $11,561, and median costs in the 2-4 year and 5-14 age groups were $5,995 and $6,233, respectively. Average costs provide a partial indication of cost differences by category due to the non-normal, positively skewed distribution of costs and wide data spread. The median (and mean) per-patient total cost of the pre-XDR-TB group was $4,632 ($13,341) compared to median (and mean) per patient total costs of $7,985 ($13,262) and $12,754 ($15,540) for the MDR/RR-TB and XDR-TB categories respectively.

**Table 2:** Median costs of course of treatment (US$) by cost category in children with multidrug-resistant/rifampicin-resistant tuberculosis, stratified by age, resistance profile, HIV status and disease site (n=271)

| Median costs (US\$)<br>(interquartile range) | Procedures | Primary healthcare<br>clinic visits | Labs | Outpatient visits | Pharmaceuticals | Hospitalisation (all) | Total |
| --- | --- | --- | --- | --- | --- | --- | --- |
| Age: |  |  |  |  |  |  |  |
| 0 to 1 years | \$42<br>(17 - 120) | \$108<br>(51 - 205) | \$288<br>(141 - 444) | \$415<br>(188 - 1,179) | \$1,216<br>(1,064 - 1,924) | \$8,654<br>(1,553 - 23,596) | \$11,561<br>(3,573 - 26,621) |
| 2 to 4 years | \$42<br>(17 - 82) | \$118<br>(61 - 205) | \$210<br>(128 - 389) | \$322<br>(188 - 509) | \$1,345<br>(1,251 - 2,423) | \$2,893<br>(0 - 22,194) | \$5,995<br>(2,955 - 24,309) |
| 5 to 14 years | \$17<br>(0 - 52) | \$205<br>(92 - 307) | \$357<br>(195 - 597) | \$295<br>(54 - 616) | \$827<br>(784 - 1,338) | \$1,645<br>(0 - 14,508) | \$6,233<br>(1,624 - 17,736) |
| Resistance profile: |  |  |  |  |  |  |  |
| MDR/RR-TB | \$34<br>(0 - 87) | \$143<br>(61 - 225) | \$274<br>(150 - 515) | \$375<br>(214 - 911) | \$1,064<br>(809 - 1,324) | \$5,274<br>(777 - 20,709) | \$7,985<br>(2,725 - 23,393) |
| PreXDR-TB | \$0<br>(0 - 52) | \$123<br>(41 - 266) | \$260<br>(128 - 415) | \$161<br>(80 - 375) | \$3,184<br>(2,683 - 3,510) | \$259<br>(0 - 25,017) | \$4,632<br>(3,628 - 28,733) |
| XDR-TB | \$52<br>(0 - 90) | \$215<br>(108 - 395) | \$494<br>(265 - 757) | \$522<br>(348 - 991) | \$4,734<br>(3,773 - 6,661) | \$5,621<br>(647 - 16,391) | \$12,754<br>(7,618 - 22,144) |
| Site of disease |  |  |  |  |  |  |  |
| Pulmonary | \$17<br>(0 - 55) | \$164<br>(72 - 266) | \$253<br>(152 - 472) | \$268<br>(107 - 509) | \$1,220<br>(812 - 2,229) | \$1,553<br>(0 - 12,361) | \$4,423<br>(2,210 - 15,374) |
| Extrapulmonary | \$52<br>(17 - 135) | \$123<br>(51 - 236) | \$341<br>(166 - 529) | \$509<br>(268 - 1,072) | \$1,167<br>(870 - 1,798) | \$10,613<br>(2,589 - 25,292) | \$14,534<br>(5,050 - 28,583) |
| HIV |  |  |  |  |  |  |  |
| HIV positive | \$17<br>(0 - 75) | \$92<br>(61 - 225) | \$505<br>(267 - 602) | \$616<br>(375 - 1,206) | \$1,037<br>(827 - 1,776) | \$17,370<br>(5,773 - 28,696) | \$20,857<br>(8,183 - 32,967) |
| HIV negative | \$34<br>(0 - 83) | \$154<br>(61 - 246) | \$258<br>(141 - 462) | \$322<br>(107 - 616) | \$1,219<br>(821 - 2,017) | \$3,149<br>(0 - 16,442) | \$6,060<br>(2,548 - 19,473) |
| All | \$33<br>(0 - 83) | \$143<br>(61 - 246) | \$286<br>(148 - 509) | \$348<br>(134 - 697) | \$1,196<br>(821 - 2,017) | \$4,401<br>(0 - 19,266) | \$7,576<br>(2,725 - 22,986) |

MDR/RR-TB = Multidrug-Resistant or Rifampicin Resistant Tuberculosis PreXDR-TB – Pre-extensively Drug-Resistant Tuberculosis; XDR-TB = Extensively Drug-Resistant Tuberculosis

**Figure 1:**
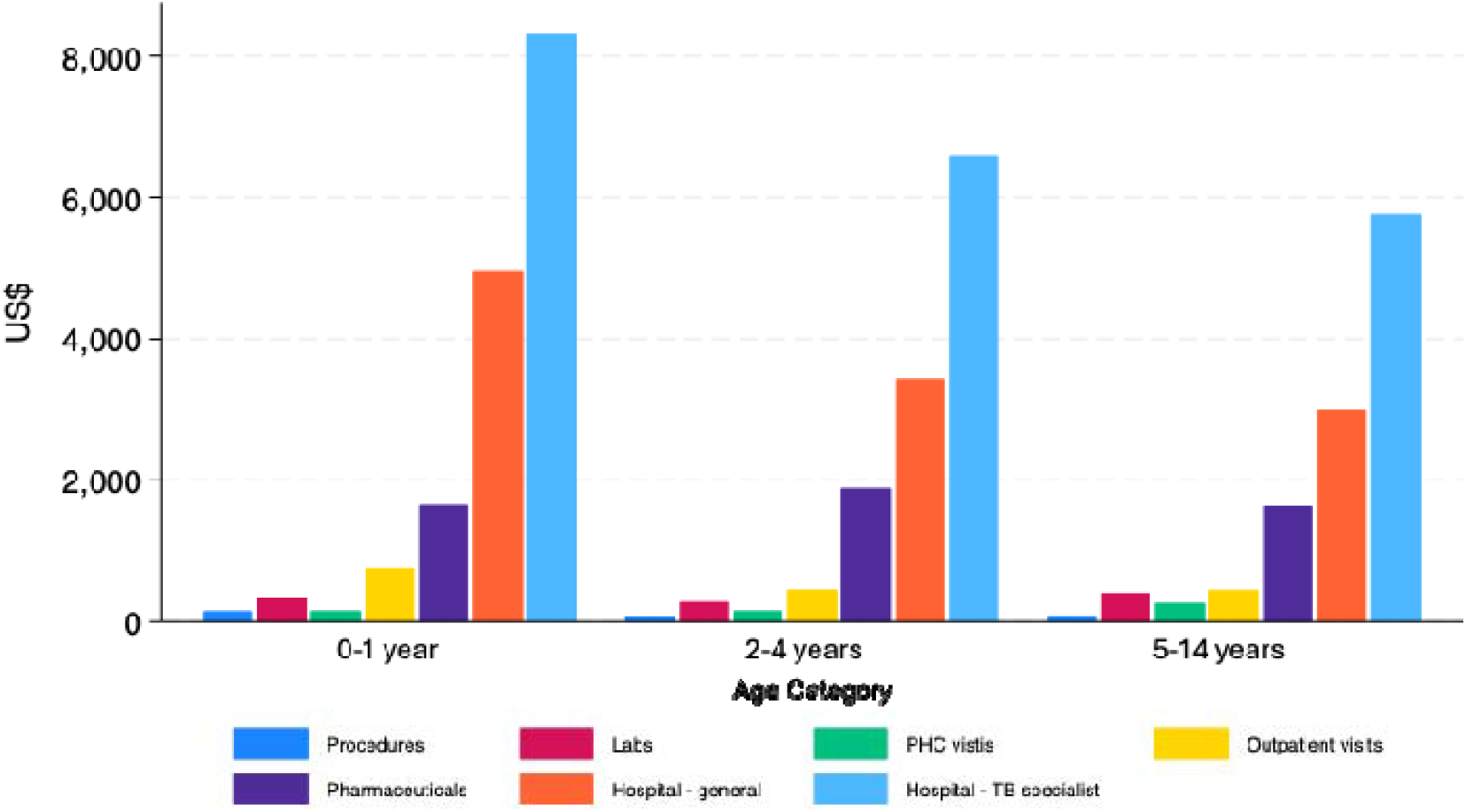
Mean costs of course of treatment (US$) by cost category in children with multidrug-resistant/rifampicin-resistant tuberculosis by age cohort (n=271)

Table 3 reflects the median total costs of treatment by age category, with stratifications by resistance profile, site of disease, sex and HIV status. The trend of higher costs for children with extrapulmonary TB and who are living with HIV was observed across age bands. While there was some variation in the highest relative median treatment cost by category between the 0-1-year-old and 2-4-year-old groups, the combined 0-4-year age group consistently had higher median total costs compared to the 5-14-year-old age group.

**Table 3:**
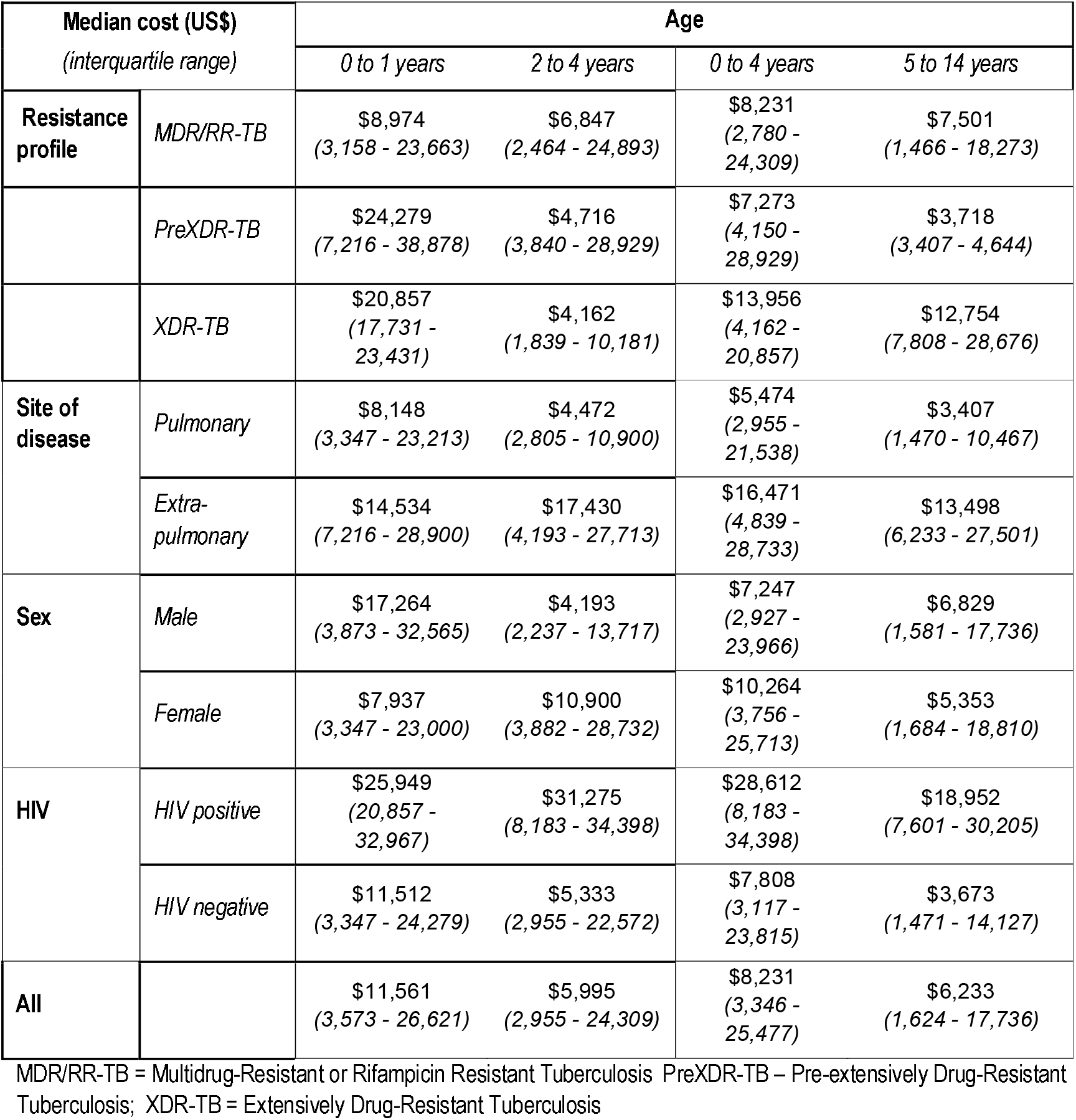
Median costs of course of treatment (US$) in children with multidrug-resistant/rifampicin-resistant tuberculosis by age category stratified by resistance profile, disease site, sex and HIV status (n=271)

| Median cost (US\$)<br>(interquartile range) | | Age | | | |
| --- | --- | --- | --- | --- | --- |
|  |  | 0 to 1 years | 2 to 4 years | 0 to 4 years | 5 to 14 years |
| Resistance profile | MDR/RR-TB | \$8,974<br>(3,158 - 23,663) | \$6,847<br>(2,464 - 24,893) | \$8,231<br>(2,780 - 24,309) | \$7,501<br>(1,466 - 18,273) |
| | PreXDR-TB | \$24,279<br>(7,216 - 38,878) | \$4,716<br>(3,840 - 28,929) | \$7,273<br>(4,150 - 28,929) | \$3,718<br>(3,407 - 4,644) |
| | XDR-TB | \$20,857<br>(17,731 - 23,431) | \$4,162<br>(1,839 - 10,181) | \$13,956<br>(4,162 - 20,857) | \$12,754<br>(7,808 - 28,676) |
| Site of disease | Pulmonary | \$8,148<br>(3,347 - 23,213) | \$4,472<br>(2,805 - 10,900) | \$5,474<br>(2,955 - 21,538) | \$3,407<br>(1,470 - 10,467) |
| | Extra-pulmonary | \$14,534<br>(7,216 - 28,900) | \$17,430<br>(4,193 - 27,713) | \$16,471<br>(4,839 - 28,733) | \$13,498<br>(6,233 - 27,501) |
| Sex | Male | \$17,264<br>(3,873 - 32,565) | \$4,193<br>(2,237 - 13,717) | \$7,247<br>(2,927 - 23,966) | \$6,829<br>(1,581 - 17,736) |
| | Female | \$7,937<br>(3,347 - 23,000) | \$10,900<br>(3,882 - 28,732) | \$10,264<br>(3,756 - 25,713) | \$5,353<br>(1,684 - 18,810) |
| HIV | HIV positive | \$25,949<br>(20,857 - 32,967) | \$31,275<br>(8,183 - 34,398) | \$28,612<br>(8,183 - 34,398) | \$18,952<br>(7,601 - 30,205) |
| | HIV negative | \$11,512<br>(3,347 - 24,279) | \$5,333<br>(2,955 - 22,572) | \$7,808<br>(3,117 - 23,815) | \$3,673<br>(1,471 - 14,127) |
| All | | \$11,561<br>(3,573 - 26,621) | \$5,995<br>(2,955 - 24,309) | \$8,231<br>(3,346 - 25,477) | \$6,233<br>(1,624 - 17,736) |

MDR/RR-TB = Multidrug-Resistant or Rifampicin Resistant Tuberculosis; PreXDR-TB – Pre-extensively Drug-Resistant Tuberculosis; XDR-TB = Extensively Drug-Resistant Tuberculosis

Figure 1 represents mean costs by category and demonstrates the high proportion of costs incurred in the under 5-year-old age group, predominantly related to general hospitalisation and specialist TB hospital stay (tabulated mean costs by category in S1 appendix). PHC clinic costs were consistently a minor element of health-service costs; PHC clinic expenditure represented just 1.5% of total per patient costs across the cohort.

Table 4 details the results of the regression analysis of log total health-service costs. The analysis demonstrated the significant independent effect of age, disease site, living with HIV, and treatment duration on total costs, providing further insight to the observed median costs in Table 2. Children living with HIV incurred 2.36 -fold higher treatment costs than those without HIV co-infection, and extrapulmonary disease was associated with a 91% increase in total costs compared to pulmonary disease, controlling for other variables. Drug resistance category did not have a significant impact on total costs in the adjusted or unadjusted analysis, despite differences in pharmaceutical costs within the categories; median pharmaceutical costs were $1,064, $3,184 and $4,734 for the MDR/RR-TB, Pre-XDR-TB and XDR-TB patients, respectively. Treatment duration was a significant driver of costs, with each additional month of treatment increasing total costs by 11% in the adjusted regression, and 12% in the unadjusted regression. In the adjusted model, cost differences were significant between age categories; with the 0-1-year-old group incurring 77% higher costs and the 2-4-year-old group incurring 59% compared to the 5-14-year-old group. In the unadjusted model, the cost differences between the 2–4- and 5-14-year-old group did not reach significance. Sex did not have a significant independent impact on total costs in the adjusted or unadjusted regression results.

**Table 4:** Regression model results – impact of patient and tuberculosis disease characteristics on log total costs of treating children with multidrug-resistant/rifampicin-resistant tuberculosis (n= 271)

| | | Adjusted<br>$\beta$ (SE) | 95% CI | Unadjusted<br>$\beta$ (SE) | (95% CI) |
| --- | --- | --- | --- | --- | --- |
| Age | 5 to 14 years (ref) |  |  |  |  |
|  | 2 to 4 years | 1.59**<br>(0.24) | 1.17 - 2.14 | 1.22<br>(0.22) | 0.85 - 1.75 |
|  | 0 to 1 years | 1.77**<br>(0.26) | 1.32 - 2.38 | 1.6**<br>(0.29) | 1.12 - 2.28 |
| Resistance profile | MDR/RR-TB (ref) |  |  |  |  |
|  | PreXDR-TB | 1.34<br>(0.22) | 0.97 - 1.84 | 1.13<br>(0.22) | 0.76 - 1.67 |
|  | XDR-TB | 1.38<br>(0.36) | 0.82 - 2.31 | 1.63<br>(0.53) | 0.86 - 3.09 |
| Site of disease | Pulmonary (ref) |  |  |  |  |
|  | Extra-pulmonary | 1.91**<br>(0.25) | 1.49 - 2.46 | 2.09**<br>(0.32) | 1.55 - 2.81 |
|  | Other/unknown | 0.69<br>(0.20) | 0.39 - 1.21 | 0.67<br>(0.22) | 0.35 - 1.3 |
| HIV status | HIV-negative (ref) |  |  |  |  |
|  | HIV-positive | 2.36**<br>(0.46) | 1.6 - 3.47 | 2.31**<br>(0.52) | 1.48 - 3.61 |
| Sex | Female (ref) |  |  |  |  |
|  | Male | 0.92<br>(0.12) | 0.72 - 1.18 | 0.98<br>(0.15) | 0.73 - 1.33 |
| Treatment duration | Months | 1.11**<br>(0.01) | 1.08 - 1.14 | 1.12**<br>(0.01) | 1.09 - 1.15 |
| | $\alpha$ | 516.25<br>(147.51) | 294.11 - 906.18 | n/a | |

\*\* P<0.01
R-squared: 0.3814
\* P<0.05
Adj. R-squared 0.3601
MDR/RR-TB = Multidrug-Resistant or Rifampicin Resistant Tuberculosis PreXDR-TB – Pre-extensively Drug-Resistant Tuberculosis; XDR-TB = Extensively Drug-Resistant Tuberculosis; SE = standard error; CI = Confidence Interval

Under the expectation that the distribution of total costs would be non-normal, a gamma distribution was estimated (see Figure 2 for the density function). The distribution of total costs yielded a mean total cost of $13,411 (95% Confidence Interval $11,789 - $15,033); further distributions by age cohorts and resistance profile are represented in S1 supplementary appendix.

**Figure 2:**
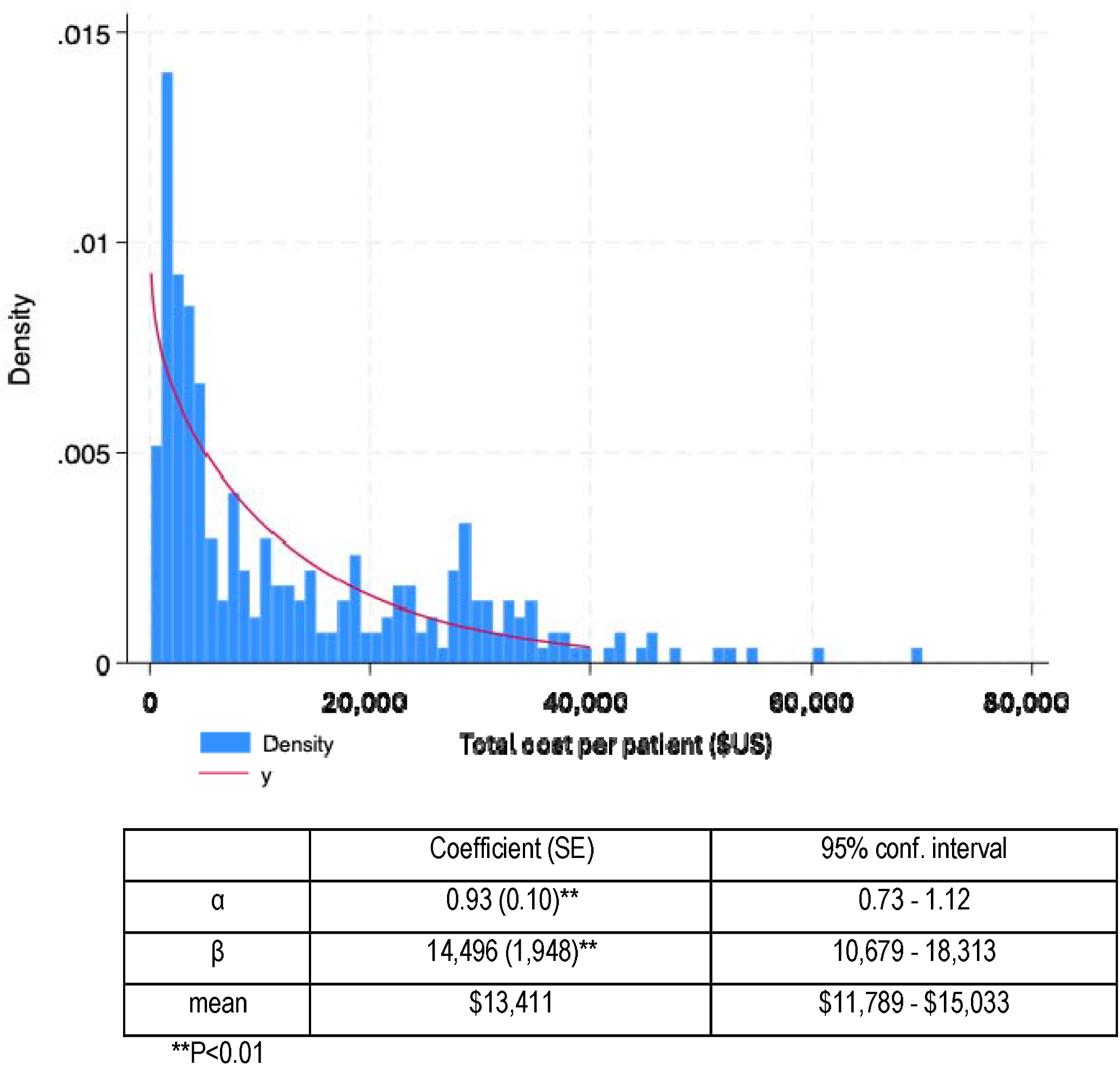
Gamma distribution of total costs of treating children aged 0-14 years for multidrug-resistant/rifampicin-resistant tuberculosis (n=271)

## DISCUSSION

This study presents a substantive empirical health-service costing analysis of childhood MDR/RR-TB in a high TB-burden, LMIC country setting, and is to our knowledge the first published empirical costs analysis specifically assessing childhood MDR/RR-TB. It provides valuable insights into the range of costs and cost drivers associated with MDR/RR-TB in children and can inform economic evaluations investigating the value for money of new treatments, vaccines, and prevention of childhood MDR/RR-TB in South Africa and other settings.

The analysis indicates that health-service costs are influenced by multiple factors demonstrating the heterogeneity of the treatment experience for children with MDR/RR-TB. Factors that increase hospital length of stay are expected to be a major determinant of costs, indicating that initiatives aimed at limiting progression to hospitalisation may yield substantial cost savings in addition to health benefits. A decentralised TB treatment model has been adopted in the Western Cape of South Africa, meaning that TB-related hospitalisation is either clinically necessary or required due to social or household conditions (18). Therefore, innovation in models of care such as increased use of community health workers, earlier access to care and social support for households are expected to have the greatest impact on costs by reducing hospitalisation. The small proportion of costs associated with PHC clinic visits indicates that there is scope to consider differentiated models of care such as home-based nursing or further community health worker support to reduce clinical indications for prolonged hospitalisation.

Recorded treatment outcome of either LTFU or treatment success was not associated with a significant difference in calculated costs. The treatment costs were not differentiated by outcome in the primary analysis, meaning that reported costs reflect actual expected health-service-costs for all patients, not only those with a successful treatment outcome. Further analysis relating to outcome is presented in supplementary appendix. The n=5 children who died had a mean total cost almost twice the sample mean cost ($23,369 vs $13,411), reflecting a high level of health care utilisation prior to death, adding to understanding that preventing severe tuberculosis disease is likely to yield significant cost savings in addition to its direct health benefits for children.

The regression analysis demonstrates the independent effect of key dimensions of disease on costs. The drug resistance profile did not have a significant cost impact, indicating that although drug regimens are generally longer and more expensive in patients with pre-XDR-TB and XDR-TB compared to MDR/RR-TB, the cost of hospitalisation outweighed drug cost differences – a child with MDR-TB who required extensive hospitalisation is expected to incur greater costs than a child with XDR-TB who requires less hospitalisation for clinical or social reasons. The cost reductions associated with adoption of decentralised models of care in adult populations were demonstrated by Sinanovic et al. (2015), where hospital-based RR-TB treatment model was compared to a community-based (decentralised) model for adult patients in the Western Cape province of South Africa (24). The analysis found the centralised model for adults was 42% more costly than the decentralised model. These findings are also supported by a comparative costs assessment of hospital vs community-based treatment models for adults with MDR-TB in the KwaZulu-Natal province in South Africa by Loveday et al. (2018) (25). The significantly higher TB treatment costs for children living with a HIV is expected to be due the increased complexity of the clinical management of co-infection and a greater likelihood of hospitalisation.

The gamma distribution enables a greater understanding of the nature and shape of costs for this cohort. The estimation of the α and β gamma coefficients will be an essential input to further analysis, particularly a comprehensive representation of cost uncertainty by enabling probabilistic sensitivity analysis. The distribution of costs demonstrates the relatively small number of patients who require very high-cost care. While the overall total median cost per patient in the cohort was US$7,567, the mean total cost was US$13,562 and the 5% highest-cost patients ranged from US38,878 to US$69,830, highlighting the limitations of utilising only sample medians and means to represent costs where the cost distribution is not normally distributed.

The analysis applies a novel approach to costing using a real-world dataset and provides an alternative to generalised cross-country costs approximations. For example, Fitzpatrick and Floyd (2012) conducted a systematic review specifically on costs and cost-effectiveness of MDR-TB treatment (26). The review identified studies only in adult populations and included no studies on the African continent; however, the authors synthesised results for the Southern African WHO region to predict costs of MDR-TB care. While the estimated results from the systematic review were reasonable approximations, the estimates were insufficient for determining local prioritisation decisions or understanding of the cost drivers and distributions. Siapka et al. (2020) conducted a global systematic review and meta-analysis of health-service costs for TB treatment in LMICs (27), noting that synthesised global cost analysis should not substitute for local, high-quality primary data. This analysis was made possible by access to real-world data through the PHDC. The PHDC is an innovative information management system, and this analysis is an example of the extensive range of benefits that can arise when digital health information is made available in a coherent and coordinated manner.

It is expected that the prior lack of costing studies in this area is due to the relatively low prevalence, long treatment duration, and case management complexity of MDR/RR-TB in children. Although this sample of 271 patients is only a proportion of the total prevalence of childhood MDR/RR-TB in South Africa, it is expected to have reasonably captured data for all patients with a final treatment outcome in the Western Cape province. This study excluded those who are lost to follow-up as it aims to represent per-patient costs of care for a completed course of treatment. Patients lost to follow-up may re-enter the system or seek care outside of the Western Cape province, therefore incurring additional costs outside of this analysis.

The incorporation of over 1.1 million data points in the initial data sample indicates that a study of this size and complexity would not be feasible with the usual manual data collection methods. Health costing analysis in high-income settings commonly use insurance and payment transaction data, while information systems based in publicly funded health settings may not comprehensively record pricing and payment data where care is free at the point of use. This study demonstrates that real-world cost analysis can be produced from routine utilisation and surveillance data and does not necessarily require collection of comprehensive health finance or pricing data. The dataset required substantive structuring and cleaning to enable the costing analysis, and as health information and database management systems expand in LMIC settings, careful consideration should be given to the structure, design, and availability of data to enable rapid production of costing analysis to more accurately inform localised priority setting and budgeting.

This analysis had some limitations, particularly linked to data availability and representation. Importantly, this analysis is limited to costs of the health provider and does not incorporate the costs incurred by households associated with accessing care.

Instances of miscoding, errors, or gaps in clinical coding may be reflected in the analysis. Gaps in areas such as medicines dispensed in PHC settings or hospital ward stock were a key area of uncertainty and were partially addressed through the estimation of expected medicine regimens for this cohort. It is expected that there was substantial activity that the “procedures” category did not capture. For example, the use of imaging including X-ray is a routine aspect of care, however 31.5% of the cohort did not have recorded procedure data, indicating that procedure costs are likely an underestimate. The analysis utilised PHDC inputs related to TB initiation, classifications, and outcomes from the national electronic TB register EDRWeb. Although this is the primary authoritative source for MDR/RR-TB surveillance data in South Africa, previous assessments have identified limitations to the use of EDRWeb data for analytical purposes (28). The drug resistance classification data extracted from the PHDC did not include indicators for whether children and adolescents had confirmed RR -TB by drug susceptibility testing or were clinically diagnosed with MDR/RR-TB based on the sensitivity of the expected adult source patient. This limited any inference related to any cost differences in patients associated with method of determining resistance profile.

Children in the preXDR-TB resistance category represented 18% of the total included cohort (Table 1), reflecting 29.5% of the children initially in the MDR-TB category (i.e. excluding those coded as RR-TB and XDR-TB). While the use of dispensing data to indicate resistance profile introduces some uncertainty, the proportion allocated to the pre-XDR-TB category using this approach aligned to a concurrent empirical drug resistance study in the Western Cape that reported the rate of fluoroquinolone resistance in children with MDR-TB was 32% (17). It is also possible that this method incorrectly allocates a patient as pre-XDR-TB who had a contraindication to fluoroquinolones, but it is expected that the risk of this incorrect classification is small.

The regression model exhibited a relatively low R^2^, (0.38, Adj R^2^ = 0.36) indicating that only a modest proportion of the variance in total health-service costs was explained by the included variables. This is not unexpected given the known heterogeneity of health-service utilisation data, which is influenced by a range of factors including clinical severity, patient and health professional behaviors and system-level factors. Although the included variables represented key TB disease characteristics, these additional factors were not captured in the dataset. The regression model did provide useful insights into important individual drivers of costs, such as age and HIV status, while confirming expectations that factors such as sex are not associated with cost differences. These insights demonstrate the value of the regression analysis, even if the overall model has low predictive power.

The data extraction period was from 2018 to 2021 and cost structures and treatment approaches for children with MDR/RR-TB are changing rapidly. The expanded use of bedaquiline for children and associated potential future adaption of shorter adult MDR/RR-TB regimen based on results of the recent TB-PRACTECAL trial (29) and BEAT-Tuberculosis trial (30), are expected to reduce costs substantially. This analysis will provide a useful reference costing at a specific point in time, against which future cost analysis can be compared.

## Data Availability

All data produced in the present study are available upon reasonable request to the authors and on approval of the Provincial Health Data Centre of the Western Cape Government Department of Health and Wellness, South Africa.

## Notes

### Competing Interest Statement

The authors have declared no competing interest.

### Funding Statement

This study did not receive any funding

