## Supplemental appendix 1 for "Health-service costs for the treatment of multidrug-resistant/rifampicin-resistant tuberculosis in children in South Africa: application of a real-world dataset"

**Manuscript Title**

### 1. Supplementary methods and inputs

#### 1.1 Costing of health facility utilisation

Patient events extracted from the PHDC included PHC clinic visits, outpatient visits, and admission and discharge from general hospitals (district, regional, tertiary) and specialist TB hospitals. The per-patient cost of health facility utilisation was calculated by multiplying the number of outpatient and PHC visits by defined “per visit” unit costs, and multiplying the length of stay at general and specialist TB hospitals by the respective “inpatient day” unit costs (Table S1). The South African National Department of Health Uniform Patient Fee Schedule (UPFS) (2021) (1) was the primary price source for the analysis and was used to calculate outpatient visit and inpatient day costs. The parameter for primary care clinic cost was sourced from literature as primary care clinic visits are not listed in the UPFS. Pooran et al 2013 (2) reported PHC clinic costs in Western Cape, the unit cost per visit was inflated to a 2021 value using the South African annual consumer price index as reported in World Bank databank (3).

Table S1: Health facility unit costs (US$) used in analysis

| **Costing component** | **Cost** | **Description** |
| --- | --- | --- |
| Clinic visit | $10.25 | Pooran et al 2013 (CPI-adjusted to 2021) |
| Outpatient visit | $23.58 | UPFS midpoint of all hospital type outpatient facility fee and medical practitioner outpatient fee |
| Hospital inpatient day: District/regional /tertiary hospital | $76.18 | UPFS inpatient facility fee and medical practitioner fee, midpoint of all hospital types, weighted for inpatient high care (20%) and inpatient chronic care (80%) |
| Hospital inpatient day: TB specialist hospital | $28.76 | UPFS hospital inpatient facility fee and daily medical practitioner and nurse fee |

#### 1.2 Costing of medicine usage and categorisation of resistance types

Medicines costs estimation was conducted using a normative approach based on National South African TB treatment guidelines (4) and clinical judgement to represent the most likely regimen by age group and drug resistance profile during the period of data collection (2018-2021) (Table S2). Costs were informed by Wilkinson et al 2024 (5) that applied weight-adjusted daily dosing price estimates by individual medicine and by regimen. Prices as reported by Wilkinson et al were adjusted to reflect the South African national medicines tender prices in 2021 (6).

Table S2. Expected medicine regimen by age and resistance classification

|  | Resistance classification | | |
| --- | --- | --- | --- |
| Age | MDR/RR-TB | Pre-XDR-TB | XDR-TB |
| 5-14 years | A | C | X |
| 2 to 4 years | B | D | X |
| 0-1 years | B | D | X |

| A | LZD (2)-BDQ(6)-hH (4-6)/ LFX-CFZ-Z-EMB (9-11) |
| --- | --- |
| B | LZD (2-6)-LFX-CFZ-TRD-PAS (12-15) |
| C | LZD (6-15)-BDQ-CFZ-TRD-DLM (15) |
| D | LZD-DLM-CFZ-TRD-PAS (15) |
| X | Regimen variable. Estimate per month drug cost as 5x weighted average cost of LZD, BDQ, PAS, TRD, DLM, CFZ for duration of treatment |

BDQ-bedaquiline, CFZ-clofazimine, DLM-delamanid, EMB-ethambutol, hH- high dose isoniazid, LFX-levofloxacin, LZD-linezolid, PAS-para-aminosalicylic acid, TRD-terizidone, , Z-pyrazinamide, RR-TB - rifampicin-resistant tuberculosis, MDR/RR-TB = Multidrug-Resistant or Rifampicin Resistant Tuberculosis; PreXDR-TB – Pre-extensively Drug-Resistant Tuberculosis; XDR-TB = Extensively Drug-Resistant-Tuberculosis

Standard-dose isoniazid may have been used under regimen A instead of high-dose isoniazid (hH) for some patients. The cost difference per month of using high-dose isoniazid or standard dose isoniazid is less than $1, therefore for the purposes of regimen cost estimation, regimen A was generalised to patients with resistance profile classified as RR-TB or MDR-TB*.*

Regimen costs as reported by Wilkinson et al 2024(5) represented weight- and aged- based dosing for linezolid oral tablets. However, in the PHDC cohort assessed in this analysis, a substantial proportion of linezolid was dispensed as an oral solution: 74% (0-1 year age group), 58% (2-4 years old age group), and 33% (5-14 year old age group). The high cost of linezolid oral solution relative to the oral tablet is expected to have a substantial impact on overall regimen costs. To adjust for this cost, regimen costs as reported by Wilkinson et al 2024 were updated with a monthly linezolid cost by age, weighted by the proportional use of the oral liquid formulation in the PHDC cohort. Patients with missing pharmaceutical data were assumed to have the same proportion of linezolid liquid use as those with pharmaceutical data. (Table S3).

Table S3. Linezolid monthly cost (US$) by formulation

|  | Cost per month | | |
| --- | --- | --- | --- |
| Age group | Linezolid 100mg/5ml oral solution | Linezolid 600mg oral tablets | Weighted monthly cost of linezolid used in analysis |
| 5-14 years | $365.09 | $8.18 | $128.78 |
| 2 to 4 years | $195.13 | $8.18 | $112.87 |
| 0-1 years | $134.18 | $8.18 | $106.18 |

The monthly medicine costs by resistance classification and age are recorded in Table S4. The cost of the applicable regimen was applied to each patient based on age category, drug resistance profile, and recorded length of treatment.

Table S4. Total regimen pharmaceutical cost (US$) by age and resistance classification

| Regimen cost for first 6 months | | | |
| --- | --- | --- | --- |
| Age group | MDR/RR-TB | Pre-XDR-TB | XDR-TB |
| 5-14 years | $721.99 | $2,176.63 | $2,521.59 |
| 2 to 4 years | $884.04 | $2,024.78 | $1,839.55 |
| 0-1 years | $776.30 | $1,814.09 | $1,645.94 |
| Regimen cost for each additional month | | | |
| Age group | MDR/RR-TB | Pre-XDR-TB | XDR-TB |
| 5-14 years | $27.49 | $210.37 | $420.27 |
| 2 to 4 years | $109.56 | $337.46 | $306.59 |
| 0-1 years | $96.06 | $302.35 | $274.32 |

MDR/RR-TB - multidrug-resistant tuberculosis or rifampicin resistant tuberculosis, pre-XDR-TB – pre extensively drug-resistant tuberculosis, XDR-TB – extensively drug-resistant tuberculosis.

#### 1.3 Costing of procedures

Procedures are recorded by patient in the PHDC dataset and include imaging (eg x-ray) and surgical interventions. For the analysis, 86 different types of procedures as reported in the PHDC were classified into categories aligning with the UPFS (Table S5). Procedures were costed by multiplying the number and type of procedure received by each patient by the UPFS price, differentiated by the type of hospital in which the procedure was performed.

Table S5. Unit cost (US$) of procedures by hospital type

| UPFS category | Unit cost  (district/regional/TB specialist hospitals) | Unit cost  (tertiary hospitals) |
| --- | --- | --- |
| Imaging A | $17 | $17 |
| Imaging B | $46 | $49 |
| Imaging D | $222 | $233 |
| Imaging E | $559 | $586 |
| Minor Theatre A | $58 | $65 |
| Minor Theatre B | $75 | $82 |
| Major Theatre C | $162 | $213 |
| Major Theatre B | $250 | $328 |
| Major Theatre C | $424 | $558 |

Note Imaging Category C is recorded in the UPFS but was not applicable to any imaging descriptors in the PHDC dataset.

#### 1.4 Costing of laboratory tests

All lab tests and investigations for the included n=271 patients were grouped into 64 categories, which consisted of various blood tests (e.g. electrolytes, blood cell counts, hormone levels), COVID investigations, HIV investigations, TB cultures, other TB diagnostic tests, drug susceptibility tests, urine tests and other minor categories. Costs were assigned to the tests falling into the 64 categories using the National Health Laboratory Service State Price List 2021 (7) (Table S6). Tests and investigations specific to TB (e.g. Xpert tests) that occurred within 5 months prior to, or after, TB treatment were included. All other tests and investigations were excluded if they occurred more than 30 days earlier than the first date of TB evidence or more than 30 days after the final TB outcome was recorded for each individual the patient

Table S6. Laboratory test classification

| NHLS cost category | Reported test | |
| --- | --- | --- |
| Bloods routine and electrolytes (n=4896) | | |
| BLOOD_ALBUMIN_PROTEIN  BLOOD_ELECTROLYTES  BLOOD_FBC_DIFF  BLOOD_TEST_OTHER  BLOOD_UREA  BLOOD_WCC_PLATELETS_ LYMPHOCYTES  CREATININE (PLUS EGFR_MDRD)  SERUM_INDICES  URINE_TEST_OTHER | ALBUMIN  BICARBONATE  CALCIUM  CHLORIDE  CREATINE KINASE (CK)  CREATININE (PLUS EGFR)  CREATININE (PLUS MDRD)  DIFFERENTIAL COUNT  FBC AND DIFF  FULL BLOOD COUNT  INORGANIC PHOSPHATE  LYMPHOCYTE SUBSETS  MAGNESIUM  PLATELET COUNT  POTASSIUM  SERUM INDICES  SODIUM | TOTAL PROTEIN  UREA  URIC ACID  URINE CALCIUM  URINE CHLORIDE  URINE CREATININE  URINE DIPSTICK  URINE INORGANIC PHOSPHATE  URINE MAGNESIUM  URINE METANEPHRINES  URINE OSMOLALITY  URINE POTASSIUM  URINE PROTEIN  URINE SODIUM  URINE UREA  WHITE CELL COUNT |
| Bloods other (n=394) | | |
| BLOOD_AMYLASE_LIPASE  BLOOD_ESR  BLOOD_FOLATE_VIT_B12  BLOOD_GLUCOSE_HBA1C  BLOOD_INR_APTT_FIBRINOGEN_ DDIMR  BLOOD_LACTATE  BLOOD_LIPIDS_CHOLESTEROL_ TRIGLYC  BLOOD_PROCALCITONIN_SEPSIS  BLOOD_RCC_HAEMOGLB_ HAEMOCRIT  BLOOD_RETICULOCYTE  BLOOD_TEST_OTHER  FERRITIN_TRANSFERRATIN_IRON  PLASMA_INDICES  SMEAR_BLOOD  TEST_IFA_FEIA_ELISA | ACETAMINOPHEN /PARACETAMOL  ACTIVATED PARTIAL THROMBOPLASTIN TIME  ADRENOCORTICOTROPIC HORMONE  ALDOSTERONE  AMYLASE  ANTI FACTOR XA ASSAY  ANTI-CCP AB (ELISA)  ANTI-DNASE B  ANTI-DSDNA AB (ELISA)  ANTI-DSDNA AB (FEIA)  ANTI-NUCLEAR AB SCREEN (FEIA)  ANTI-NUCLEAR FACTOR (IFA)  ANTI-RNP AB (ELISA)  ANTI-SM AB (ELISA)  ANTI-SMOOTH MUSCLE AB (IFA)  ANTI-SS-A (RO) AB (ELISA)  ANTI-SS-B (LA) AB (ELISA)  ANTI-STREPTOLYSIN O TITRE (AUTOMATED)  ANTI-THYROID AB (HAEMAGGLUTINATION)  BETA-HCG  CANCA: ANTI-PROTEINASE 3 AB (ELISA)  CANCA: ANTI-PROTEINASE 3 AB (FEIA)  CARBAMAZEPINE / TEGRETOL  CHOLINESTERASE  COMPLEMENT C3  COMPLEMENT C4  CORTISOL  ECHINOCOCCUS (ELISA)  EDTA PLASMA INDICES  ERYTHROCYTE SEDIMENTATION RATE  FERRITIN | FIBRINOGEN  GLUCOSE (FASTING)  GLUCOSE (RANDOM)  GLYCATED HAEMOGLOBIN (HBA1C)  HAEMOGLOBIN  HAPTOGLOBIN  INDIRECT COOMBS  INT NORMALISED RATIO (INR)  IRON  LACTATE  LACTATE DEHYDROGENASE (LD)  LIPASE / LIPID PROFILE  LUTEINISING HORMONE (LH)  LYMPHOCYTE SUBSETS  MEAN CELL VOLUME  MICRODELETION SYNDROMES  OSMOLALITY  OXIDATIVE BURST  PANCA: ANTI-MYELOPEROXIDASE AB (ELISA)  PANCA: ANTI-MYELOPEROXIDASE AB (FEIA)  PERIPHERAL BLOOD MORPHOLOGY  PHENOBARBITAL  PROCALCITONIN (SENSITIVE)  RENIN  RETICULOCYTE COUNT  RHEUMATOID FACTOR  SERUM FOLATE  THYROGLOBULIN  TOTAL CHOLESTEROL  TRANSFERRIN  TRIGLYCERIDE  VALPROATE / EPILIM  VITAMIN B12 |
| COVID (n=73) | | |
| COVID_ANTIGEN  COVID_PCR | CLINIC COVID-19 RAPID AG (PANBIO)  CSA: COVID-19 RAPID AG (RAPIGEN)  CSA: COVID-19 RAPID AG (STANDARDQ) | SARS-COV-2 PCR  SARS-COV-2 RAPID AG  SARS-COV-2 RT-PCR  SARS-COV-2 TOTAL AB (ROCHE) |
| Diagnostics (n=3479) | | |
| CULTURE_PUS_CSF_URINE_STOOL_BLOOD  MENINGITIS_PCR  RESP_VIRUS_PCR  STAIN_AFB  STAIN_AURAMINE  STAIN_AURAMINE_W_TBCULTURE  STAIN_GRAM_ALL  STAIN_OTHER  TB ANTIGEN  TB_CULTURE | ACID FAST STAIN FOR PARASITES  AFB (POSITIVE MGIT CULTURE)  AURAMINE (WITH TB CULTURE)  AURAMINE O STAIN  AUTOMATED CULTURE  CALCAFLUOR STAIN FOR FUNGI  CULTURE CATHETER TIP  CULTURE FOR CSF  CULTURE ID BACTERIAL  CULTURE MYCOLOGY  CULTURE PUS  CULTURE RESP/SPUTUM  CULTURE STERILE SITES  CULTURE STOOL  CULTURE TISSUE  CULTURE URINE  CYTOLOGY SPECIAL STAINS  GRAM STAIN CSF | GRAM STAIN PUS  GRAM STAIN RESP/SPUTUM  GRAM STAIN STERILE SITES  GRAM STAIN TISSUE  MENINGITIS SCREEN PCR  NICD: AFB (POSITIVE CULTURE)  NICD: ATYPICAL PNEUMONIA PCR  NICD: DST (POSITIVE CULTURE)  NICD: TB CULTURE LM  NICD: TB CULTURE MIDDLEBROOK  NICD: TB MP-ANTIGEN ID  NICD: TB+  RESP VIRUSES MULTIPLEX PCR  SPUTUM GRAM STAIN (BARTLETTS)  TB BLOOD CULTURE  TB CULTURE LIQUID MEDIUM  TB MP-ANTIGEN ID  ZIEHL-NEELSEN STAIN |
| Diagnostics GeneXpert (n=552) | | |
| GENEXPERT | GENEXPERT | GENEXPERT ULTRA |
| Drug sensitivity (n=418) | | |
| LAB_PATH_SERVICE_OTHER  LPA FIRST LINE  LPA SECOND LINE  MGIT SENSITIVITY (1ST LINE)  MGIT SENSITIVITY (2ND LINE) | MGIT SENSITIVITY (1ST LINE)  MGIT SENSITIVITY (2ND LINE)  NICD: DRUG SENSITIVITY TESTING - PYRAZINAMIDE  NICD: MGIT SENSITIVITY (2ND LINE)  NICD: MGIT SENSITIVITY (XTENDED) | PCR LINE PROBE (CM)  TB LPA FIRST LINE (CLIN SAMPLE)  TB LPA FIRST LINE (ISOLATE)  TB LPA SECOND LINE (CLIN SAMPLE)  TB LPA SECOND LINE (ISOLATE)  TB SENSITIVITY 1LD (AGAR) |
| Infections other (n=279) | | |
| ANTIGEN_TEST_OTHER  BLOOD_CRP  BLOOD_IMMUNOGLOBULIN  BLOOD_TEST_OTHER  BLOOD_TPALLIDUM_RPR_FTA_ SYPHILIS  CARAPENEM_REST_SCREEN  CMV_OTHER _VIROLOGY_ANTIGEN  CMV_PCR  HEPATITIS ANTIGEN | C-REACTIVE PROTEIN  C.DIFF AG AND TOXIN TEST  CMV QUALITATIVE PCR  CMV VIRAL LOAD  CRE SCREEN  CRYPTOCOCCAL AG (LFA) REFLEX  CRYPTOCOCCAL ANTIGEN LFA  EPSTEIN BARR VIRUS VCA IGM  FTA (SYPHILIS)  HEP B SURFACE AB  HEP B SURFACE AG  HEPATITIS A IGM  HEPATITIS B CORE IGM | HEPATITIS B CORE TOTAL AB  HEPATITIS C ANTIBODY  HLA-B27  IMMUNOGLOBULIN A  IMMUNOGLOBULIN G  IMMUNOGLOBULIN M  RPR (SYPHILIS)  RUBELLA IGG  RUBELLA IGM  T.PALLIDUM ANTIBODIES  TOTAL IGE  TOXOPLASMA IGM ELISA  URINE LEGIONELLA ANTIGEN TEST |
| Liver function (n=912) | | |
| LIVER_FUNCTION_TESTS | ALANINE TRANSAMINASE (ALT)  ALKALINE PHOSPHATASE (ALP)  ASPARTATE TRANSAMINASE (AST)  CONJUGATED BILIRUBIN | GAMMA-GLUTAMYL TRANSFERASE (GGT)  NEONATAL TOTAL BILIRUBIN  TOTAL BILIRUBIN |
| Thyroid (n=722) | | |
| BLOOD_TSH_PTH_T3_T4 | FOLLICLE STIMULATING HORMONE (FSH)  PARATHYROID HORMONE (PTH)  THYROID STIMULATING HORMONE (TSH) | THYROXINE (FREE T4)  TRI-IODO THYRONINE (FREE T3) |
| Other (n=227) | | |
| CSF_CELL_COUNT_INDIA_INK  CSF_CHLORIDE_GLUCOSE_PROTEIN_ADENOSINE  FAECAL_OTHER  LAB_PATH_SERVICE_OTHER  LAB_TEST_UNKNOWN  URINE_CELL_COUNT  BLOOD_TEST_OTHER | 25-OH VITAMIN D (IMMUNOASSAY)  CSF ADENOSINE DEAMINASE  CSF CELL COUNT  CSF CELL COUNT (AUTOMATED)  CSF CHLORIDE  CSF GLUCOSE  CSF IDENTIFICATION OF FLUID  CSF PROTEIN  FAECAL CALPROTECTIN  FLUID ADENOSINE DEAMINASE  FLUID ALBUMIN  FLUID CYTOSPIN | FLUID GLUCOSE  FLUID LD  FLUID PROTEIN  HISTOLOGY (GSH)  HISTOLOGY (TBH)  HISTOLOGY INTERNAL REFERRAL  INDIA INK  LAB TEST_UNKNOWN  NICD: RPOB SEQUENCING  URINE CELL COUNT/HPF  URINE CELL COUNT/ML |

### 2. Supplementary results

#### 2.1 Mean costs of treatment

Table S7. Mean costs of course of treatment (US$) by cost category in children with multidrug-resistant/rifampicin-resistant tuberculosis, stratified by age, resistance profile, HIV status and disease site (n=271)

| Mean costs ($US)  (standard deviation) | Procedures | Primary healthcare clinic visits | Labs | Outpatient visits | Pharmaceuticals | Hospitalisation (all) | Total |
| --- | --- | --- | --- | --- | --- | --- | --- |
| Age |  |  |  |  |  |  |  |
| 0 to 1 years | $149 (326) | $151 (138) | $334 (248) | $756 (842) | $1,644 (1,151) | $13,254 (13,294) | $16,288 (14,439) |
| 2 to 4 years | $85 (147) | $152 (160) | $278 (221) | $450 (414) | $1,883 (1,133) | $10,022 (12,964) | $12,870 (13,306) |
| 5 to 14 years | $87 (344) | $258 (331) | $408 (267) | $446 (465) | $1,631 (1,787) | $8,750 (12,052) | $11,580 (12,782) |
| Resistance profile |  |  |  |  |  |  |  |
| MDR/RR-TB | $126 (334) | $182 (205) | $346 (255) | $589 (634) | $1,104 (525) | $10,915 (12,798) | $13,262 (13,606) |
| PreXDR-TB | $31 (46) | $188 (191) | $301 (211) | $311 (504) | $3,151 (1,041) | $9,360 (13,671) | $13,341 (14,261) |
| XDR-TB | $72 (95) | $379 (590) | $514 (303) | $660 (454) | $5,057 (2,476) | $8,860 (10,304) | $15,541 (11,178) |
| Site of disease |  |  |  |  |  |  |  |
| Pulmonary | $56 (129) | $222 (295) | $334 (248) | $418 (482) | $1,775 (1,555) | $7,588 (11,021) | $10,392 (11,698) |
| Extrapulmonary | $187 (436) | $167 (159) | $375 (254) | $741 (725) | $1,661 (1,292) | $14,521 (13,233) | $17,654 (14,040) |
| HIV |  |  |  |  |  |  |  |
| HIV positive | $191 (599) | $167 (182) | $471 (238) | $828 (599) | $1,645 (1,345) | $20,006 (17,701) | $23,308 (18,397) |
| HIV negative | $94 (223) | $198 (252) | $330 (252) | $504 (604) | $1,716 (1,451) | $9,196 (11,413) | $12,038 (12,179) |
| All | $106 (295) | $195 (244) | $348 (254) | $543 (611) | $1,707 (1,436) | $10,512 (12,807) | $13,411 (13,563) |

MDR/RR-TB = Multidrug-Resistant or Rifampicin Resistant Tuberculosis; PreXDR-TB – Pre-extensively Drug-Resistant Tuberculosis; XDR-TB = Extensively Drug-Resistant-Tuberculosis

Note: the very high reported standard deviation relative to the mean indicates the very wide spread (positive skewed) consistent with the fitted gamma distribution.

#### 2.2 Cost distributions:

##### a. Cost distribution by Age 0-1

Figure F1: Gamma distribution of total costs (US$) for age group years 0-1 (n =84)

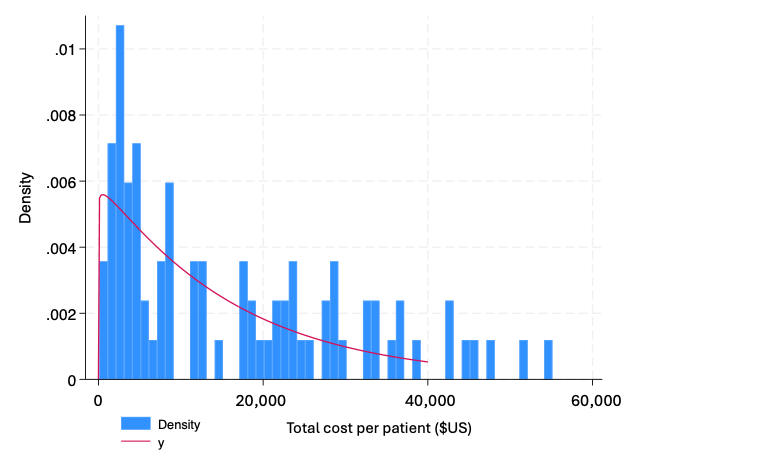

Table F1 – Distribution parameters of total costs for age group years 0-1 (n=84)

|  | Coefficient (SE) | 95% conf. interval |
| --- | --- | --- |
| α | 1.03 (0.18)** | 0.68 - 1.39 |
| β | 15,773 (3,852)** | 8,223 - 23,323 |
| mean | $16,288 (1,575) | $13,154 - $19,421 |

**P<0.01

##### b. Cost distribution by Age 2-4

Figure 2: Gamma distribution of total costs (US$) for age group years 2-4 (n =78)

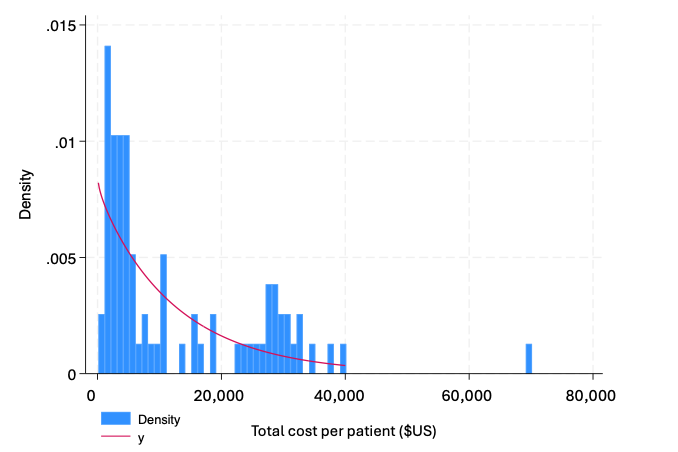

Table F2 – Distribution parameters of total costs (US$) for age group years 2-4 (n=78)

|  | Coefficient (SE) | 95% conf. interval |
| --- | --- | --- |
| α | 0.98 (0.20)** | 0.59 - 1.36 |
| β | 13,159 (3,050)** | 7,180 - 19,138 |
| mean | $12,870 (1,507) | 9,870 - 15,870 |

**P<0.01

##### c. Cost distribution by Age 0-4

Figure 3: Gamma distribution of total costs (US$) for age group years 0-4 (n = 162)

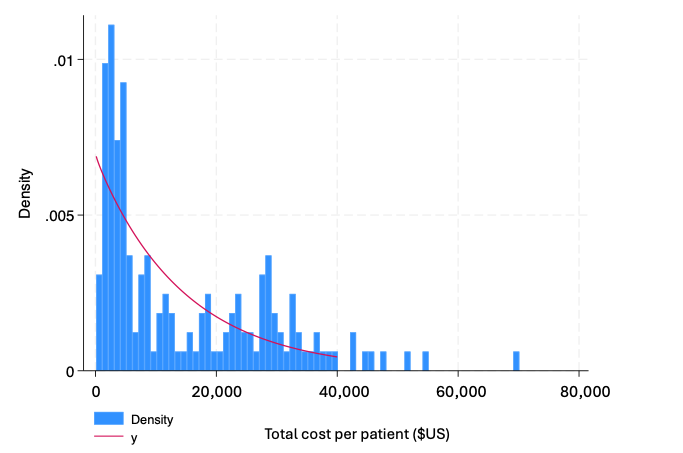

Table F3 – Distribution parameters of total costs (US$) for age group years 0-4 (n =162)

|  | Coefficient (SE) | 95% conf. interval |
| --- | --- | --- |
| α | 0.99 (0.13)** | 0.73 - 1.25 |
| β | 14,725 (2,517)** | 9,793 - 19,658 |
| mean | $14,642 (1,097) | 12,475 - 16,809 |

**P<0.01

##### d. Cost distribution by Age 5-14

Figure 4: Gamma distribution of total costs (US$) for age group years 5-14 (n = 109)

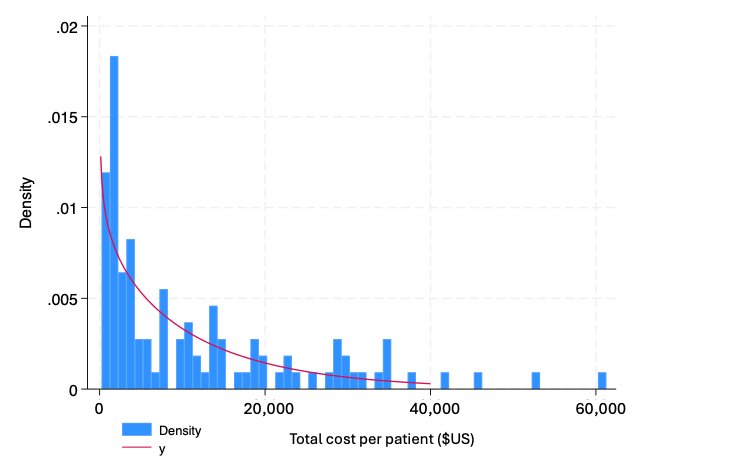

Table F4 – Distribution parameters of total costs (US$) for age group years 5-14 (n 109)

|  | Coefficient (SE) | 95% conf. interval |
| --- | --- | --- |
| α | 0.86 (0.16)** | 0.55 - 1.17 |
| β | 13,513 (2,859)** | 7,910 - 19,115 |
| mean | $11,580 (1,224) | 9,154 - 14,007 |

**P<0.01

##### e. Cost distribution by drug resistance classification – MDR/RR-TB

Figure 5: Gamma distribution of total costs (US$) for children classified as either MDR or RR-TB (n =206)

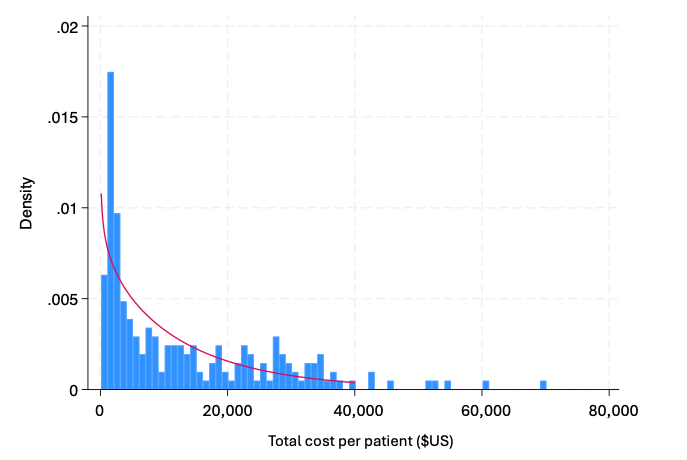

Table F5 – Distribution parameters of total costs (US$) for children classified as either RR-TB or MDR-TB (n = 206)

|  | Coefficient (SE) | 95% conf. interval |
| --- | --- | --- |
| α | 0.88 (0.11)** | 0.67 - 1.09 |
| β | 15,131 (2,346)** | 10,532 - 19,730 |
| mean | $13,262 (0,948) | 11,393 - 15,131 |

**P<0.01

Limited numbers of patients in the pre-XDR-TB and XDR-TB resistance profiles limited reliable specification of cost distributions for these sub-groups.

#### 2.3 Stepwise assessment of regression model

A stepwise modelling strategy was used to inform the regression model (Model 1). The model specification was informed by forward selection and backward elimination methods with parameter selection based on model fit and requirement for the model to test theoretical understanding of factors that drive cost structures for treating children with tuberculosis. The use of interaction terms for Age, HIV, Site and Resistance categories were rejected from the model due to limited improvements in overall model fit and interpretation. The stepwise assessment involved the sequential addition of predictor variables to the model allowing the assessment of the incremental contribution of each variable to the overall model fit. The results of the stepwise assessment are detailed in Table S8.

Length of treatment (treatment months) had the largest explanatory value within the model, (adj. R^2^ = 0.233), with subsequent valuables providing only small improvements in the model fit. Notably, resistance profile and sex had very modest improvements on model fit. However, these remain important for the theoretical interpretation of the regression model for understanding factors that do (and do not) have a meaningful difference on the cost structure or treatment children with MDR/RR-TB.

*Model 1*:

$Log \left( Total costs \right)_{i} = \alpha+ \beta_{1}{Age}_{ij} + \beta_{2}{Resistance}_{ik} + \beta_{3}{Disease\_Site}_{is} {+ \beta}_{4}{Sex}_{i} + \beta_{5}{HIV}_{i}+ \beta_{6}{Trmt\_Duration}_{i}+ \varepsilon_{i}$

Table S8. Stepwise Regression impact of patient and tuberculosis disease characteristics on log total costs of treating children with multidrug-resistant/rifampicin-resistant tuberculosis (n= 271)

| Stepwise regression model  β (SE) | | 1 | 2 | 3 | 4 | 5 | 6 |
| --- | --- | --- | --- | --- | --- | --- | --- |
| Treatment duration | Months | 1.12** (0.01) | 1.12** (0.01) | 1.11** (0.01) | 1.11** (0.01) | 1.11** (0.01) | 1.11** (0.01) |
| Site of disease | *Pulmonary (ref)* |  |  |  |  |  |  |
|  | Extra-pulmonary |  | 1.91** (0.26) | 1.93** (0.25) | 1.87** (0.24) | 1.9** (0.24) | 1.91** (0.25) |
|  | Other/unknown |  | 0.84 (0.25) | 0.7 (0.2) | 0.68 (0.19) | 0.69 (0.2) | 0.69 (0.2) |
| HIV status | *HIV-negative (ref)* |  |  |  |  |  |  |
|  | HIV-positive |  |  | 2.00** (0.39) | 2.36** (0.46) | 2.40** (0.47) | 2.36** (0.46) |
| Age | *5 to 14 years (ref)* |  |  |  |  |  |  |
|  | 2 to 4 years |  |  |  | 1.58** (0.24) | 1.57** (0.24) | 1.59** (0.24) |
|  | 0 to 1 years |  |  |  | 1.71** (0.25) | 1.77** (0.26) | 1.77** (0.26) |
| Resistance profile | *MDR/RR-TB (ref)* |  |  |  |  |  |  |
|  | PreXDR-TB |  |  |  |  | 1.35 (0.22) | 1.34 (0.22) |
|  | XDR-TB |  |  |  |  | 1.38 (0.36) | 1.38 (0.36) |
| Sex | *Female (ref)* |  |  |  |  |  |  |
|  | Male |  |  |  |  |  | 0.92 (0.12) |
|  | α | 2,122 (318) | 1,778 (272) | 881 (219) | 544 (148) | 491 (135) | 516 (148) |
|  | N | 271 | 271 | 271 | 271 | 271 | 271 |
|  | R^2^ | 0.233 | 0.301 | 0.333 | 0.369 | 0.38 | 0.381 |
|  | adj. R^2^ | 0.23 | 0.293 | 0.322 | 0.355 | 0.361 | 0.36 |

^*^ p < 0.05; ^**^ p < 0.01

MDR/RR-TB = Multidrug-Resistant or Rifampicin Resistant Tuberculosis PreXDR-TB – Pre-extensively Drug-Resistant Tuberculosis; XDR-TB = Extensively Drug-Resistant Tuberculosis; SE = Standard Error

#### 2.4 Additional analysis: hospital length of stay

Hospital length of stay was a major determinant of costs; hospital stay representing 78% of total treatment costs across the cohort (81%, 78% and 76% of total costs in the 0-1 year, 2-4 year, and 5-14 year group respectively). Hospital length of stay was also highly skewed: 70 (26%) of patients recorded no hospitalisation, and the 50^th^, 75^th^ and 95^th^ percentile length of stay was 20, 161, and 315 days of hospitalisation respectively (skewness/kurtosis tests for normality = Pr(skewness 0.000, Pr(kurtosis 0.7204), n = 271). Increased hospital length of stay had a weak positive correlation with total treatment duration (Pearson’s correlation coefficient = 0.338). As a direct calculated input to the cost of treatment, hospital length of stay was not incorporated into the regression model of log total costs. However, summary statistics were generated to inform further analysis. Aligning to findings associated with Total Treatment Costs, higher mean length of stay was observed in children in younger age cohorts, extra-pulmonary TB, and children who were diagnosed as HIV positive (Table S9).

The direct relationship between length of stay and treatment costs indicates that understanding of the main drivers of long hospital stay is a large part of understanding costs of treatment. In a separate study cited in the main text, analysis of paediatric admissions to a specialist TB hospital in the Western Cape demonstrated that reasons for admission are often multidimensional and include clinical severity, site of disease, resistance profile, and social/caregiver related (8). The available data for this analysis does not enable inference of the reasons for admission and length of hospital stay, and is expected to be the principal reason for a relatively low regression model fit.

Table S9. Length of hospital stay (in days) by patient and disease characteristics (n= 271)

| Variable |  | Mean Length of Stay (days) |
| --- | --- | --- |
| Age | 0 to 1 years | 109.7 |
|  | 2 to 4 years | 85.1 |
|  | 5 to 14 years | 74.4 |
| Resistance profile | MDR/RR-TB | 87.3 |
|  | PreXDR-TB | 95.9 |
|  | XDR-TB | 80.4 |
| Site of disease | Pulmonary | 65.4 |
|  | Extrapulmonary | 122.1 |
| HIV status | HIV positive | 154.4 |
|  | HIV negative | 79.3 |
| Sex | Male | 87.5 |
|  | Female | 89.2 |
| All |  | 88.4 |

MDR/RR-TB = Multidrug-Resistant or Rifampicin Resistant Tuberculosis PreXDR-TB – Pre-extensively Drug-resistant tuberculosis; XDR-TB = Extensively Drug-Resistant Tuberculosis

A binary categorisation was applied where “hospitalisation – low” was applied to cost data where the child had less than 10 days in hospital across the entire treatment period, and “hospitalisation – high” where the child had 10 or more days in hospital. It is acknowledged that this categorisation has the limitations of a contrived proxy measure, however it will facilitate secondary analysis that can reflect scenarios involving hospital length of stay differences. The median and mean costs associated with this categorisation are shown in Table S10.

Table S10. Mean and median total costs (US$) by Hospitalisation utilisation category and age (n=271)

| Age | Hospitalisation - high | | | Hospitalisation – low | | |
| --- | --- | --- | --- | --- | --- | --- |
|  | N | Mean (SD) | Median (IQR) | N | Mean (SD) | Median (IQR) |
| 0-1 years | 58 | $22,433 (13,369) | $21,357 (21,053) | 26 | $2,578 (1,255) | $2,485 (1,427) |
| 2-4 years | 42 | $21,332 (13,118) | $23,269 (19,203) | 36 | $2,999 (1,339) | $2,880 (2,184) |
| 0-4 years | 100 | $21,971 (13,208) | $22,055 (19,299) | 62 | $2,822 (1,311) | $2,708 (1,877) |
| 5-15 years | 51 | $21,664 (12,356) | $18,952 (17,586) | 58 | $2,714 (2,160) | $1,789 (2,328) |
| All | 151 | $21,867 (12,886) | $20,185 (18,746) | 120 | $2,770 (1,766) | $2,327 (2,219) |

Low hospitalisation = length of stay <10 days total

High hospitalisation = length of stay >=10 days total

SD = Standard Deviation

#### 2.5 Additional analysis: patient outcomes

Included patients had recorded outcome classification of either “Treatment Success” (n=178), “Lost to Follow Up” (LTFU) (n = 87), Died (n=5), or “Unsuccessful Treatment” (n= 1), (Table S11). There were multiple instances of patients who were recorded as LTFU but where there was health service utilisation recorded in the dataset after the LTFU entry date, indicating that there are likely some instances of miscoding of the LTFU outcome. The principal cost analysis and regression was conducted without stratification by outcome, meaning that costs in the main analysis reflect actual costs incurred by the health system for treatment of childhood MDR-RR-TB, rather than only those with recorded completed treatment. For comprehensiveness, the Outcome classification was explored in supplementary analysis: Table S11 represents the patient and disease characteristics by Outcome, and Table S12 represents total costs stratified by Outcome. Outcome classification was not associated with any significant differences in distribution by patient and disease characteristics, but patients who were classified as LTFU had a significantly shorter treatment duration (means 11.3 vs 9.0 months). This did not result in any significant differences in Total costs between the LTFU and Treatment Success groups.

Table S11. Children with multidrug-resistant/rifampicin-resistant tuberculosis demographic and clinical characteristics by age category (n=271)

|  | Treatment success | LTFU | Died | All | Fisher’s exact  p-value |
| --- | --- | --- | --- | --- | --- |
| Characteristics | n (%^†^) | n (%^†^) | n (%^†^) | n (%^‡^) |  |
| Total in outcome category^‡^ | 178 | 87 | 5 | 271 |  |
| Female | 95  (53%) | 51  (59%) | 4  (80%) | 151  (56%) | *0.449* |
| Age category |  |  |  |  | *0.820* |
| 0-1y | 51  (61%) | 31  (37%) | 2  (2%) | 84  (31%) |  |
| 2-4y | 55  (71%) | 22  (28%) | 1  (1%) | 78  (29%) |  |
| 5-15y | 72  (66%) | 34  (31%) | 2  (2%) | 109  (40%) |  |
| Resistance profile |  |  |  |  | *0.199* |
| MDR/RR-TB | 137  (77%) | 65  (75%) | 4  (80%) | 206  (76%) |  |
| PreXDR-TB | 33  (19%) | 15  (17%) | 1  (20%) | 49  (18%) |  |
| XDR-TB | 8  (4%) | 7  (8%) | 0  (0%) | 16  (6%) |  |
| Site of TB disease |  |  |  |  | *0.967* |
| Pulmonary TB | 97  (54%) | 51  (59%) | 3  (60%) | 152  (56%) |  |
| Extrapulmonary TB | 72  (40%) | 31  (36%) | 2  (40%) | 105  (39%) |  |
| Other / Unknown | 9  (5%) | 5  (6%) | 0  (0%) | 14  (5%) |  |
| HIV + | 20  (11%) | 12  (14%) | 1  (20%) | 33  (12%) | *0.548* |
| Mean number of months of treatment (SD)** | 11.3  (4.4) | 9.0  (6.3) | 9.7  (5.7) | 10.5  (5.2) |  |

MDR/RR-TB = Multidrug-Resistant or Rifampicin Resistant Tuberculosis; PreXDR-TB – Pre-extensively Drug-Resistant Tuberculosis; XDR-TB = Extensively Drug-Resistant-Tuberculosis

**Months of treatment by Treatment Success vs LTFU Mann-Whitney U test statistic: 10.76; z = -3.281, p = 0.001

† = of total number in the applicable age category. ‡ = of total cohort (n=271) “Unsuccessful treatment” category not included in table above, reflecting N=1 child (characteristics: Age 5-14 years; XDR-TB, Pulmonary TB, HIV-).

Table S12. Total costs of course of treatment (US$) by Outcome in children with multidrug-resistant/rifampicin-resistant tuberculosis, stratified by age, sex, resistance profile, disease site and HIV status (n=271)

| Characteristic | Recorded Outcome | | | |  |  |
| --- | --- | --- | --- | --- | --- | --- |
|  | Treatment success | LTFU | Died | All | Mann-Whitney U test^†^ | |
| Age |  |  |  |  | Z score | p |
| 0-1y | $16,053  (13,277) | $15,259  (15,755) | $38,197  (6,962) | $16,288  (14,439) | -0.531 | 0.5956 |
| 2-4y | $13,015  (13,610) | $11,779  (12,622) | $28,929  (0) | $12,870  (13,306) | -0.541 | 0.5883 |
| 5-15y | $11,498  (12,783) | $12,400  (13,276) | $5,762  (5,463) | $11,580  (12,782) | 0.467 | 0.6405 |
| Sex |  |  |  |  |  |  |
| Female | $13,784  (13,007) | $11,941  (12,178) | $20,893  (18,675) | $13,267  (12,873) | -0.889 | 0.3742 |
| Male | $12,686  (13,564) | $15,133  (16,191) | $33,274  (0) | $13,592  (14,437) | -0.889 | 0.3742 |
| Resistance profile |  |  |  |  |  |  |
| MDR/RR-TB | $13,373  (13,456) | $12,491  (13,610) | $21,980  (19,410) | $13,262  (13,606) | -0.545 | 0.5858 |
| PreXDR-TB | $12,292  (13,661) | $14,612  (15,781) | $28,929  (0) | $13,341  (14,261) | -0.323 | 0.7471 |
| XDR-TB | $15,586  (7,279) | $17,531  (14,558) | $0  (0) | $15,541  (11,178) | -0.116 | 0.9079 |
| Site of TB disease |  |  |  |  |  |  |
| Pulmonary TB | $9,658  (9,650) | $11,508  (14,408) | $18,215  (21,911) | $10,392  (11,698) | -0.385 | 0.700 |
| Extrapulmonary TB | $17,604  (14,550) | $16,903  (13,013) | $31,101  (3,073) | $17,654  (14,040) | 0.216 | 0.8292 |
| Other / Unknown | $17,576  (23,146) | $8,569  (12,918) | $0  (0) | $14,360  (20,027) | -1.000 | 0.3173 |
| HIV status |  |  |  |  |  |  |
| HIV + | $25,600  (18,872) | $20,628  (18,305) | $9,625  (0) | $23,308  (18,397) | -0.856 | 0.3918 |
| HIV - | $11,712  (11,524) | $12,083  (12,926) | $26,805  (17,633) | $12,038  (12,179) | -0.15 | 0.8809 |
| Total | $13,272  (13,243) | $13,262  (13,979) | $23,369  (17,095) | $13,411  (13,563) | -0.258 | 0.7966 |

^†^ Mann-Whitney U test of Total Cost by Treatment Success vs LTFU outcome by respective category (due to small n in outcome catergory “Died”).

MDR/RR-TB = Multidrug-Resistant or Rifampicin Resistant Tuberculosis; PreXDR-TB – Pre-extensively Drug-Resistant Tuberculosis; XDR-TB = Extensively Drug-Resistant-Tuberculosis

#### 2.6 Additional analysis: patient categorisation

While this is primarily a costing analysis, further summary statistics were developed to provide additional insights into the distribution of the cohort between categories. Tables below represent proportion of the sample by resistance profile that were classified as either extra-pulmonary TB (Table S13) and HIV positive (Table S14). Neither of the comparisons found a significant proportion of patients that were classified to a specific resistance profile, confirming the expectation of limited proportional trend between resistance profile, site of disease, and HIV status.

Table S13. Proportion of cohort classified as Extra-Pulmonary Tuberculosis by resistance profile

| **Resistance profile** | **Proportion** |
| --- | --- |
| MDR/RR-TB (n=206) | 40.3% |
| Pre-XDR-TB (n=49) | 32.7% |
| XDR-TB (n=16) | 37.5% |
| All (n=271) | 38.7% |
| Pearson chi-square = 1.984 (p = 0.739) | |

MDR/RR-TB = Multidrug-Resistant or Rifampicin Resistant Tuberculosis; PreXDR-TB – Pre-extensively Drug-Resistant Tuberculosis; XDR-TB = Extensively Drug-Resistant-Tuberculosis

Table S14. Proportion of cohort classified as HIV positive by resistance profile

| **Resistance profile** | **Proportion** |
| --- | --- |
| MDR/RR-TB (n=206) | 12.6% |
| Pre-XDR-TB (n=49) | 10.2% |
| XDR-TB (n=16) | 12.5% |
| All (n=271) | 12.2% |
| Pearson chi-square = 0.2179 (p = 0.897) | |

MDR/RR-TB = Multidrug-Resistant or Rifampicin Resistant Tuberculosis; PreXDR-TB – Pre-extensively Drug-Resistant Tuberculosis; XDR-TB = Extensively Drug-Resistant-Tuberculosis
